# Identification of Spatiotemporal Associations of Social Determinants of Health on the Incidence of Adverse Birth Outcomes in Louisiana

**DOI:** 10.64898/2026.04.06.26349198

**Authors:** José Irizarry-Ayala, Jian Li, W Susan Cheng, David R. Crosslin

**Affiliations:** Tulane University, New Orleans, LA

## Abstract

**Introduction:** Louisiana ranks last in the United States of America in terms of maternal health outcomes. Previous works have highlighted the impact of some social determinants of health on the incidence of adverse birth outcomes. These works have subjectively selected specific social determinants of health from larger datasets. Here, we attempt to replicate their results with objective variable selection techniques.

**Methods:** By deriving principal components from the Agency of Healthcare Research and Quality’s parish-level social determinants of health dataset, we were able to objectively find social determinants of health associations instead of the conventional subjective variable selection approach. Then, we applied Bayesian linear mixed-effects models to calculate more conservative parameter estimates about the effects of social determinants of health on adverse birth outcome incidence. Then, we used local Moran’s I to identify clusters of spatially autocorrelated parishes. Finally, we combined the results of these two methods and inspected the relationship between important predictors and clusters of spatial autocorrelation.

**Results:** We identified several significant effects on the incidence of adverse birth outcomes, including populational composition and economic attainment, and several clusters of high and low incidences of adverse birth outcomes in Louisiana. There was also a concordant relationship between important predictors from our predictive models and the cluster assignments of Local Moran’s I.

**Conclusion:** Our results validate previous works in the subject area and hold implications for precision development of maternal health interventions in Louisiana.

## Introduction

Adverse Birth Outcomes (ABO) is an umbrella term for conditions or pathological processes associated with pregnancy. They can happen during or after pregnancy and include but are not limited to low birthweight (LBW), preterm birth (PTB), maternal or infant mortality, post-term birth, or pre-eclampsia. It is well documented that child-bearing persons living in the Southeastern United States are especially susceptible to ABO.[1] To exemplify this, Louisiana ranked last among all US states and Puerto Rico in terms of maternal and child health outcomes.[1] All these factors compound and result in Louisiana having one of the highest rates for ABO across the territorial U.S. Some work has been conducted into elucidating which factors affect rates of ABO. [2–7] One group of factors that are thought to heavily influence the incidence of ABOs are the Social Determinants of Health (SDoH). SDoH are the non-biological social factors which affect health outcomes. They reflect the societal conditions people are exposed to throughout their life course. When SDoH disproportionately affect a health outcome it is considered a health disparity. In short, health disparities are a particular type of health difference that is closely linked with economic, social, or environmental disadvantage.[8] Previous health disparities research indicates African American and/or Black people and people with low Socioeconomic Status (SES) are more likely to suffer ABO.[2, 6] These works have been essential in understanding the effects of SDoH on ABO incidence. One potential limitation of these works which we sought to address was the subjective selection of variables from larger SDoH datasets. We expect an analysis which objectively selects variables from such a dataset will replicate these previously documented associations. Another important factor which influences the rates of incidence of health outcomes like ABO is geographical location. The relationship between geography and health can combine multiple factors including local politics, environmental exposure, and proximity to resources.[9, 10] Historically, spatial factors such as high temperature and environmental contamination have been found to be related to ABO incidence.[11, 12] The passage of time can also modify the rates of incidence of a disease particularly by way of representing a change in populations, policy, access to resources, or weather events.[9] Measuring the difference in rates of ABOs can show whether over time, health disparities related to their incidence have reduced or increased.

Here, we sought to answer if the incidence of ABO varies by geography and/or over time, what social factors affect rates of ABO more generally and how heterogeneous the distribution of ABO is across Louisiana. To measure the spatiotemporal associations of ABO in Louisiana we used Bayesian statistical methods to find the distribution of the effect sizes of both spatial and temporal factors. Because we also wanted to measure the effects of social factors, we used a statistical method which allowed us to best estimate spatiotemporal associations and the effects of SDoH. Our technique of choice for this was Bayesian linear mixed-effects (BLME) models. These not only allow us to group our observations in terms of year and location, but also to estimate the effects of SDoH on ABO incidence rates. Another analytical method we applied here is local Moran’s I.[13] We used local Moran’s I to provide another view on the spatial associations of ABO rates in Louisiana. Local Moran’s I allows us to identify patterns of spatial correlation between geographies. Through examining those correlational patterns, we can identify clusters of high or low incidence of ABOs. Finally, we combined the results of both modelling approaches to inspect the relationships between the best predictors from our BLME models and the cluster labels assigned by Local Moran’s I. We are the first group to apply both Bayesian linear mixed-effects models and Local Moran’s I to the explanation of differences and identification of clusters related to ABO in Louisiana.

## Methods

### Data

To answer our three proposed questions, we combined data from various sources. Specifically, we recovered ABO data from the Louisiana Department of Health’s data portal.[6] We used the percentage of LBW and PTB at the parish-level, the Louisiana equivalent of counties, from the years 2009 to 2020 as our outcomes of interest. LBW, as defined by the Louisiana Department of Health, is a birth with a weight less than 2,500 grams. Similarly, PTB was defined as a birth before 37 weeks of gestation. These make up the only birth outcomes data that are publicly available through the Louisiana Department of Health’s portal which is reported on a yearly level. Other ABOs of interest could be maternal mortality or infant mortality. That said, maternal mortality is only publicly available on a state reporting level, eliminating the possibility of identification of parish-level associations. Furthermore, infant mortality is reported on a parish level but only in 5-year intervals, eliminating sensitivity in the detection of temporal associations. Notably, the ABO data we used is standardized by populational size. Since parishes with high populations would assuredly have higher absolute counts of ABO and vice versa, normalizing by population size ensures a fairer comparison between parishes. All parishes had at least one year of data available.

To capture social factors, we used the SDoH data from the Agency of Healthcare Research and Quality (AHRQ) from 2009 to 2020. These data contain measurements of SDoH variables from several government agencies and collection instruments. For example, total population as measured by the census or Gini index from the American Community Survey. Initially, throughout all years there were 1330 different variables with varying degrees of missingness. To address this limitation and prepare our data for modeling, we first subset the larger dataset to only include variables which had less than 30% missing observations and were numeric (n=667). In addition to data missingness, another notable limitation to consider is the expected correlational patterns between the data from the AHRQ. For example, while the correlation would probably not be perfect, we expect geographies with higher population size to have more health practitioners, households, people between the ages of 0 and 17, and any other populational variable related to number of people. While the information and subsequent interpretation that they provide is different, most statistical modelling approaches cannot discern useful information from these interdependent variables by themselves. Therefore, to best use the AHRQ dataset, we had to reduce the dimensionality of the dataset while still representing the information well.

We dropped one additional variable which contained a reported low birth weight rate by parish. It is also important to note that the variable available in the AHRQ’s data was not concordant with the data from the Louisiana Department of Health. We decided to continue using the data from the Louisiana Department of Health because it is a primary source of this information. Afterwards, we imputed the missing values. Our imputation was carried out by replacing missing values for each variable with the median value of the non-missing observations of that same parish. In the rare case that no observations for a particular variable were available for a parish, we imputed their values with the median value from across all parishes. In terms of our dependent variables, we had 116 missing observations for LBW and 7 missing observations for PTB. We handled these missing values by imputing them based on the median value of their respective parish, similarly to our independent variables. This ensured we did not have to drop any observations from our models due to missing values

Using this newly imputed dataset, we computed principal components. By calculating the principal components of our dataset, we were able to derive variables that best captured the information from the larger dataset while minimizing the correlation between them. This allows us to better satisfy the assumption of independence between our input variables. We selected how many principal components to use based on the percentage of variance explained by each principal component.

### Analysis Methods

To quantify the effects of location, time and SDoH on the incidence of ABOs in Louisiana we analyzed our data using BLME models. As mentioned previously, BLME models allow for the parameter estimation of both fixed and random effects. Fixed effects are equivalent to conventional linear regression coefficients. These represent between-group variation, or factors that are distributed differently between different groups. On the other hand, random effects, sometimes referred to as grouping variables, represent within-group variation. That is, factors that affect the sampling of other factors. For these reasons, in our model specification, we selected parish and year as random effects. We then specified a linear combination of a set of selected principal components as fixed effects. With this format, we fit two BLME models, one for LBW and the other for PTB, using the R package *rstanarm* for R version 4.4.3.[14] Otherwise, we utilized default parameters for our models. This automatically calculates a weakly informative prior for each variable. We checked for model convergence using the R-hat statistic, and assessed model performance by comparing the posterior predictive distributions to the true distributions of our outcomes as well as with Bayesian R-squared.[15] We characterized the impacts of our input variables by looking at the magnitude and 95% Confidence Region (CR) for our regression coefficients.

We calculated local Moran’s I statistics[13] for each parish using the R package rgeodat for R version 4.4.3.[16] The relevant equations for the calculation of local Moran’s I can be seen in Equations 1 and 2. Plainly, local Moran’s I compares the value of a variable at an individual locations, in this case the parishes of Louisiana, to the mean value of that same variable across all locations in the data set. It also compares that value to the neighboring geographies and determines whether those values are concordant or discordant with their neighbors. In that way it classifies geographies as high or low in terms of their relation to the whole area, as well as high or low when compared to their neighbors. For example, a geography with a significantly higher than average value for a variable surrounded by other geographies with higher-than-average values would be classified as a high-high geography. To calculate local Moran’s I we used the queen neighbors setting as opposed to the rook neighbor setting, which is more permissive in assigning geographies as neighbors. We visually inspected clusters with their respective z-score and p-value.

Then, to synthesize the results from the BLME models and local Moran’s I, we performed an integrated analysis. We selected the most important predictors, as identified by our model for each of our two outcomes, transformed them according to their respective coefficient estimate, and projected them into principal component space. Consequently, we overlayed each observation with their local Moran’s I assigned cluster label. Through this we were able to visually inspect the similarities between geographies assigned certain cluster labels and our SDoH-derived principal components as well as hypothesize latent predictors which were not considered within the input dataset.

## Results

### Variable Missingness, Imputation, and Dimensionality Reduction

We generated a stacked histogram to better visualize data missingness by variable (Figure 1). After selecting the 667 variables which had a total missingness of less than 70%, we imputed missing values as previously described. The distributions of our outcomes can be seen in Figure 2 while their spatial distribution can be seen in Figure 3. Due to the highly intercorrelated nature of our dataset, we looked to reduce the total dimensionality using principal component analysis. The scree plot resulting from our principal component analysis can be seen in Figure 4. We evaluated our principal components based on the percentage of variance explained by each one. We selected the first 9 principal components since this allows us to capture over 50% of the variance in the dataset while still greatly reducing the amount of input variables used in our Bayesian Linear Mixed Effects Models. Reducing the dimensionality of our data was of high importance because of how much it allowed us to reduce the complexity of our BLME models. The complexity of BLME models increases exponentially with each input variable added, making models with many input variables very computationally expensive. A complete look at variable loadings for our principal components can be seen in Supplemental Figure 1. A summarized version of the PC loadings can be seen in Table 1.

**Table 1.** Top 10 variables and qualitative labels for first 9 principal components derived from AHRQ SDoH data.

| PC | Variance Explained (%) | Top 10 Variables | Qualitative Label |
| --- | --- | --- | --- |
| 1 | 27 | ACS_TOT_HU, CCBP_ANNUAL_TOT_POP, ACS_TOT_HH, ACS_TOT_POP_WT, ACS_TOT_POP_US_ABOVE1, CDCW_TOT_POPULATION, ACS_TOT_POP_ABOVE_15, ACS_TOT_POP_ABOVE16, ACS_TOT_CIVIL_NONINST_POP, ACS_TOT_POP_POV | Populational size and density |
| 2 | 9.8 | SAIPE_PCT_POV_0_17, ACS_PCT_PERSON_IN_BELOW99, SAIPE_PCT_POV_5_17, ACS_MEDIAN_HH_INC, ACS_PCT_PERSON_INC_ABOVE200, SAIPE_PCT_POV, ACS_PCT_NONVET_POV_18_64, ACS_MEDIAN_INC_M, ACS_PCT_HH_INC_10000, ACS_PCT_HH_INC_99999 | Economic demographics |
| 3 | 4.4 | MP_PCT_ADVTG_PEN, MGVS_PCT_BEN_FFS_WHITE, CDCA_HEART_HOSP_RATE_ABOVE65, ACS_PCT_VET, CDCAP_HIVDIAG_RATE_ABOVE13, ACS_PCT_WHIT_NONHISP, NEPTHN_TEMPERATURE_90, ACS_PCT_NVR_MARRIED_F, ACS_PCT_NVR_MARRIED_M, MGVS_PCT_BEN_FFS_BLACK | Racioethnic demographics |
| 4 | 3.1 | ACS_PCT_AGE_ABOVE65, ACS_MEDIAN_AGE, ACS_MEDIAN_AGE_FEMALE, ACS_PCT_HH_ABOVE65, ACS_MEDIAN_AGE_MALE, WUSTL_AVG_PMG25, ACS_PCT_AGE_0_4, NOAAC_MIN_TEMP_JUN, ACS_PCT_HU_UTILITY_GAS, ACS_PCT_AGE_50_64 | Age breakdown |
| 5 | 2.9 | SAHIE_PCT_UNINSURED64_138FPL, NOAAC_MAX_TEMP_JUN, SAHIE_PCT_UNINSURED64_200FPL, SAHIE_PCT_UNINSURED64_250FPL, NEPTHN_TEMPERATURE_95, WUSTL_AVG_PM25, NEPTHN_TEMPERATURE_90, SAHIE_PCT_UNINSURED64_400FPL, NEPTHN_TEMPERATURE_100, ACS_PCT_POV_WHITE | Weather conditions |
| 6 | 2.7 | ACS_PCT_AGE_18_44, ACS_PCT_AGE_45_64, ACS_PCT_AGE_18_29, ACS_MEDIAN_AGE, ACS_MEDIAN_AGE_MALE, ACS_PCT_AGE_50_64, ACS_MEDIAN_AGE_FEMALE, ACS_PCT_VET_HS, CCBP_SOGS_RATE, ACS_PCT_POSTHS_ED | Age breakdown |
| 7 | 2.6 | ACS_PCT_FEMALE, ACS_PCT_MALE, ACS_PCT_WHITE_FEMALE, ACS_PCT_WHITE_MALE, ACS_PCT_BLACK_FEMALE, ACS_PCT_BLACK_MALE, ACS_PCT_ADMIN, ACS_PCT_DIF_STATE, ACS_PCT_GRP_QRT, ACS_PCT_PVT_PROFIT | Gender breakdown |
| 8 | 1.9 | AHRF_IP_DAY_NH_HOSP_RATE, AHRF_HOSP_BED_RATE, AHRF_TOT_HOSP_BEDS_LT, AHRF_IP_DAY_STNG_LT_RATE, AHRF_HOSP_BEDS_LT_RATE, CCD_TOT_EXPENDITURE, CCD_TOT_REVENUE, CCD_TOT_LOCAL_REVENUE, LTC_AVG_OBS_SUCCESSFUL_DISC_RATE, AHRF_OP_VST_LT_ER_OP_RATE, | Healthcare utilization |
| 9 | 1.5 | POS_SNF_RATE, ACS_PCT_DISABLE, ACS_PCT_HU_PLUMBING, ACS_PCT_NONVET_DISABLE_18_64, ACS_PCT_DRIVE_2WORK, POS_HHA_RATE, MGVS_PER_CAPITA_ACTUAL_HC, MGVS_PER_CAPITA_STD_HC, ACS_PCT_TAXICAB_2WORK, ACS_PCT_HU_KITCHEN | Transportation |

**Figure 1.**
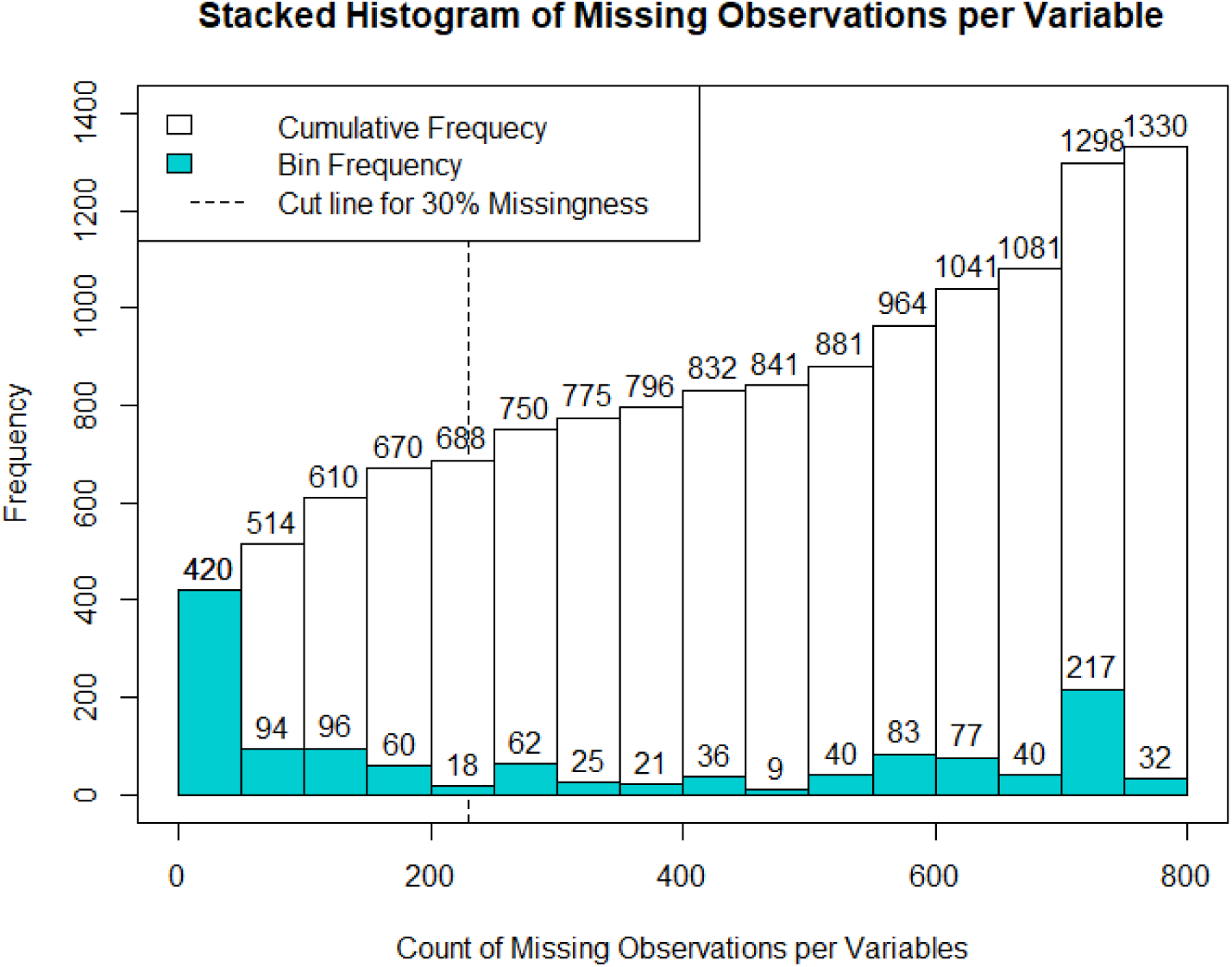
Stacked histogram of the count of missing values per variable. The blue histogram in the front represents the number of variables that are only within that bin. The white histogram in the back represents the cumulative number of variables in that bin. The vertical dashed line represents the cut line for variables that have more than 30% missing observations before filtering out non-numeric variables.

**Figure 2.**
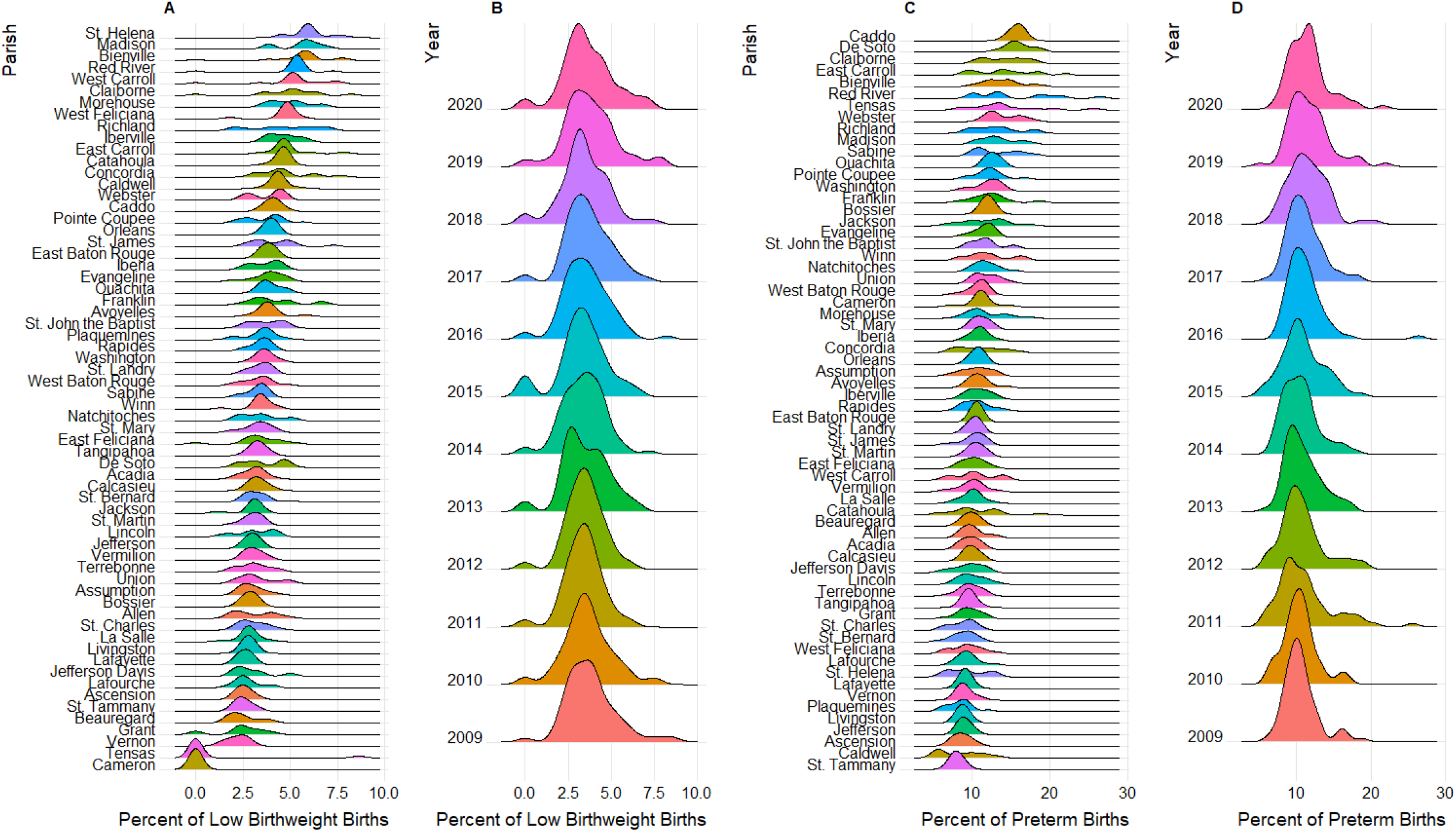
Ridgeline plots of the distributions of LBW and PTB. A and C are stratified by parish, where each year is an observation. B and D are stratified by year, where the 64 parishes make up the observations.

**Figure 3.**
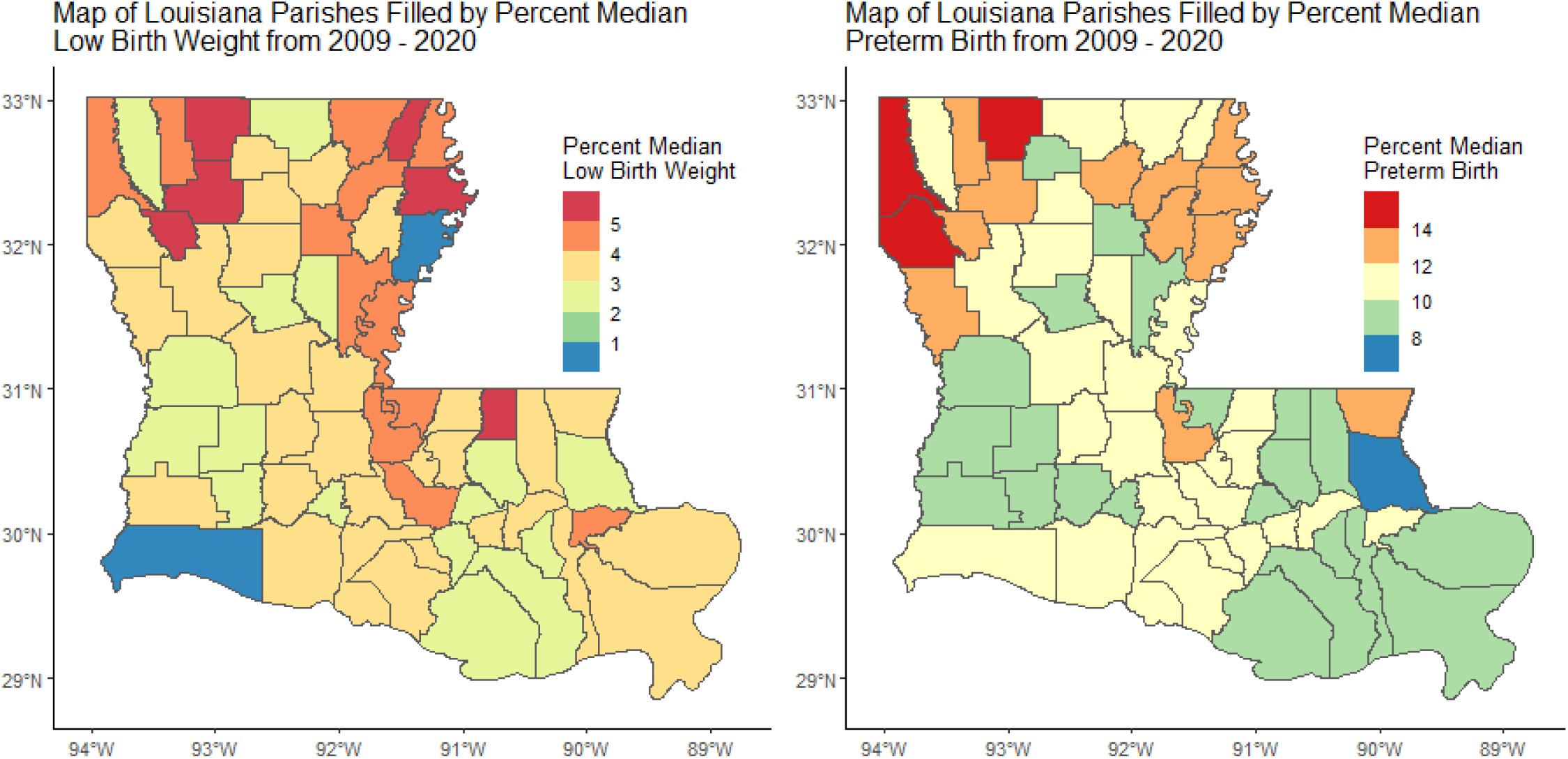
Chloropleth maps of median percentage LBW and PTB, respectively by Louisiana parish. The median was calculated from the distribution of each Parish shown in Figure 2.

**Figure 4.**
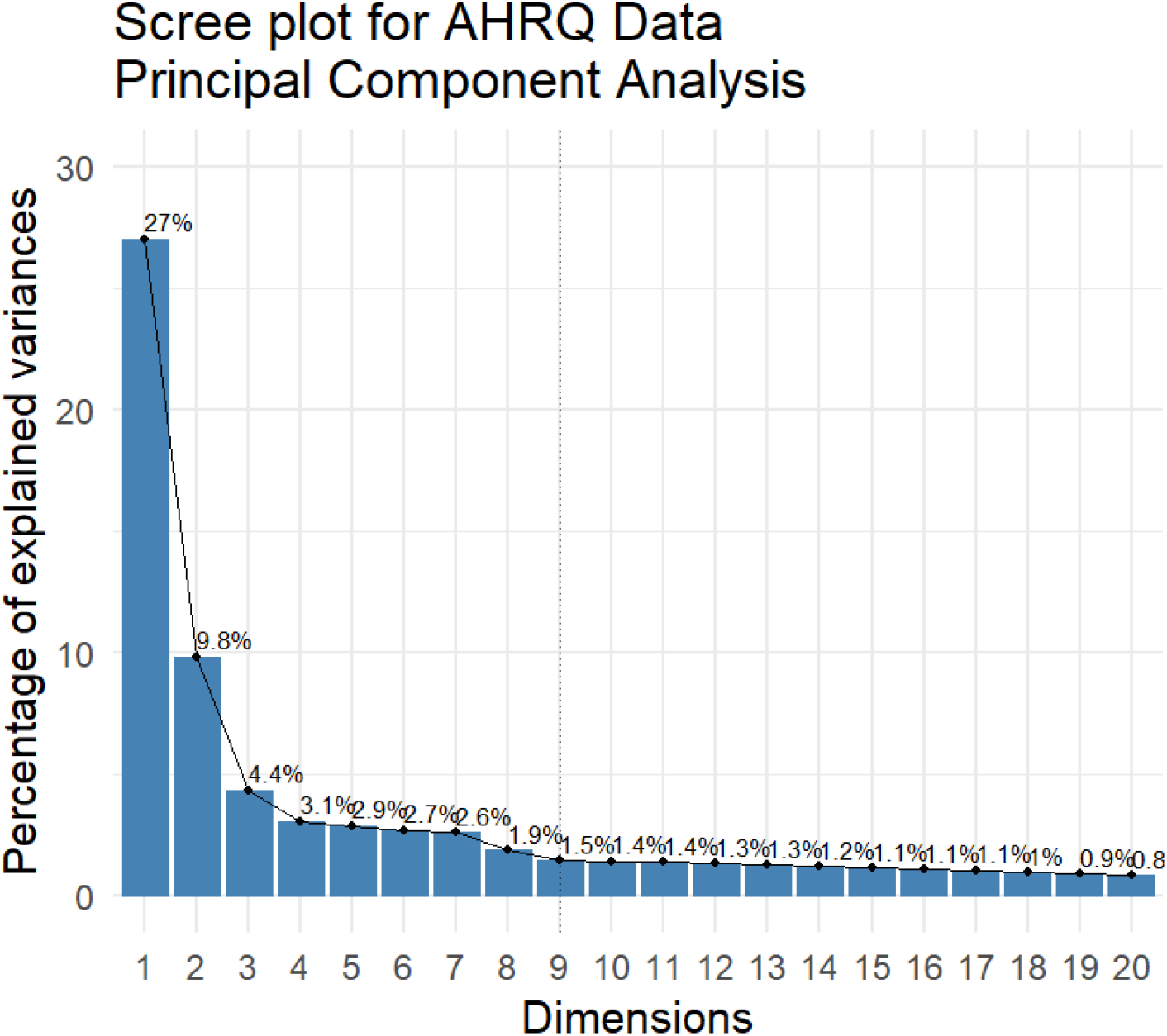
Scree plot of the principal component analysis resulting from the imputed subset of the AHRQ data. A dotted line is present on principal component 9 to show which principal components we selected for analysis.

### Bayesian Linear Mixed-Effects Model

Our models fit our data with an R-hat statistic of 1 for all parameters. The R-hat statistics indicate our models converged successfully. We further assessed model fit by comparing the data’s true distribution with the predicted distributions for our models (Figure 5). Finally, the median Bayesian R-squared for LBW was 52.05% (47.39%-56.21%) and 46.07% (40.95%-50.80%) for PTB. In combination, our mixed-effects models identified several significant predictors for PTB and LBW (Table 2, Figure 6). For LBW the significant predictors were the intercept, PC 2, PC 3, as well as several parishes: St. Helena, Bienville, West Carroll, Red River, Plaquemines, Morehouse, Richland, Caldwell, Catahoula, Jackson, Lincoln, Grant, East Feliciana, Cameron and Tensas. For PTB the significant predictors were the intercept, PC1, PC 2, and PC 8 in addition to the following parishes: De Soto, Caddo, Lincoln, Orleans, St. Helena, and Caldwell. From this, our models indicate that social determinants of health are, generally, predictors that are correlated with the incidence of ABOs in Louisiana. To best interpret which social determinants are most associated with the incidence of ABOs we must break down our significant principal components in terms of the variables that contribute the most to the difference between principal component dimensions. To accomplish this, we examined the correlation circles (Figure 8) between the significant principal components. Examining these circles revealed patterns of which variables were most associated with each outcome and the directionality of their relationships. Both outcomes were found to be significantly associated with economic attainment and the racioethnic composition of the parish. LBW, specifically, was also associated with population density and healthcare utilization.

**Table 2.**
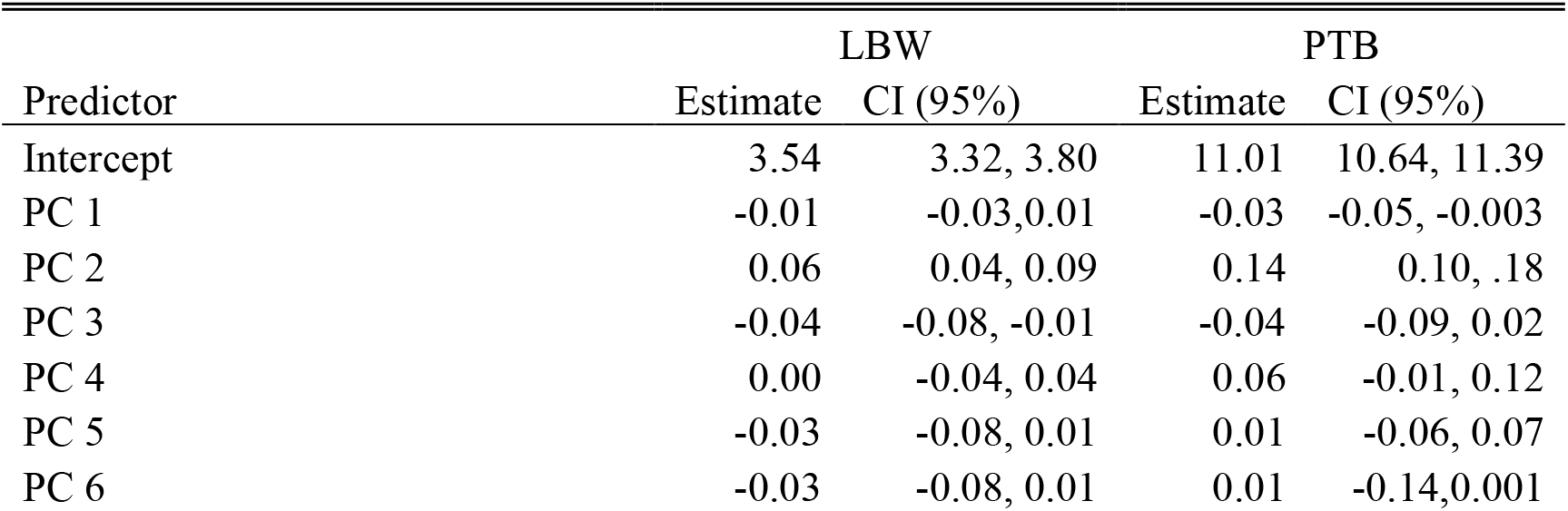
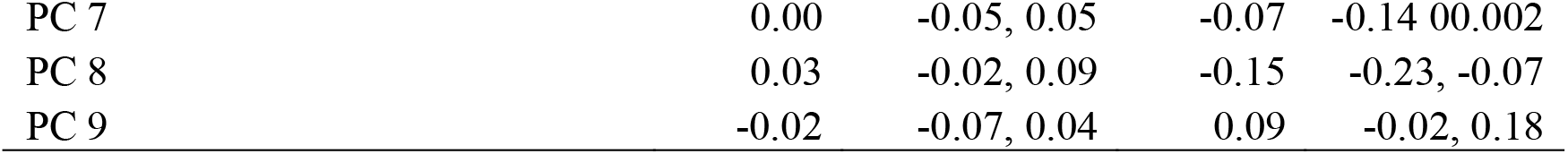
Fixed effects regression coefficients and their 95% confidence intervals for LBW and PTB. For LBW, the significant fixed effects were the intercept, PC 2, and PC 3. For PTB the significant fixed effects were the intercept, PC1, PC 2, and PC 8.

**Figure 5.**
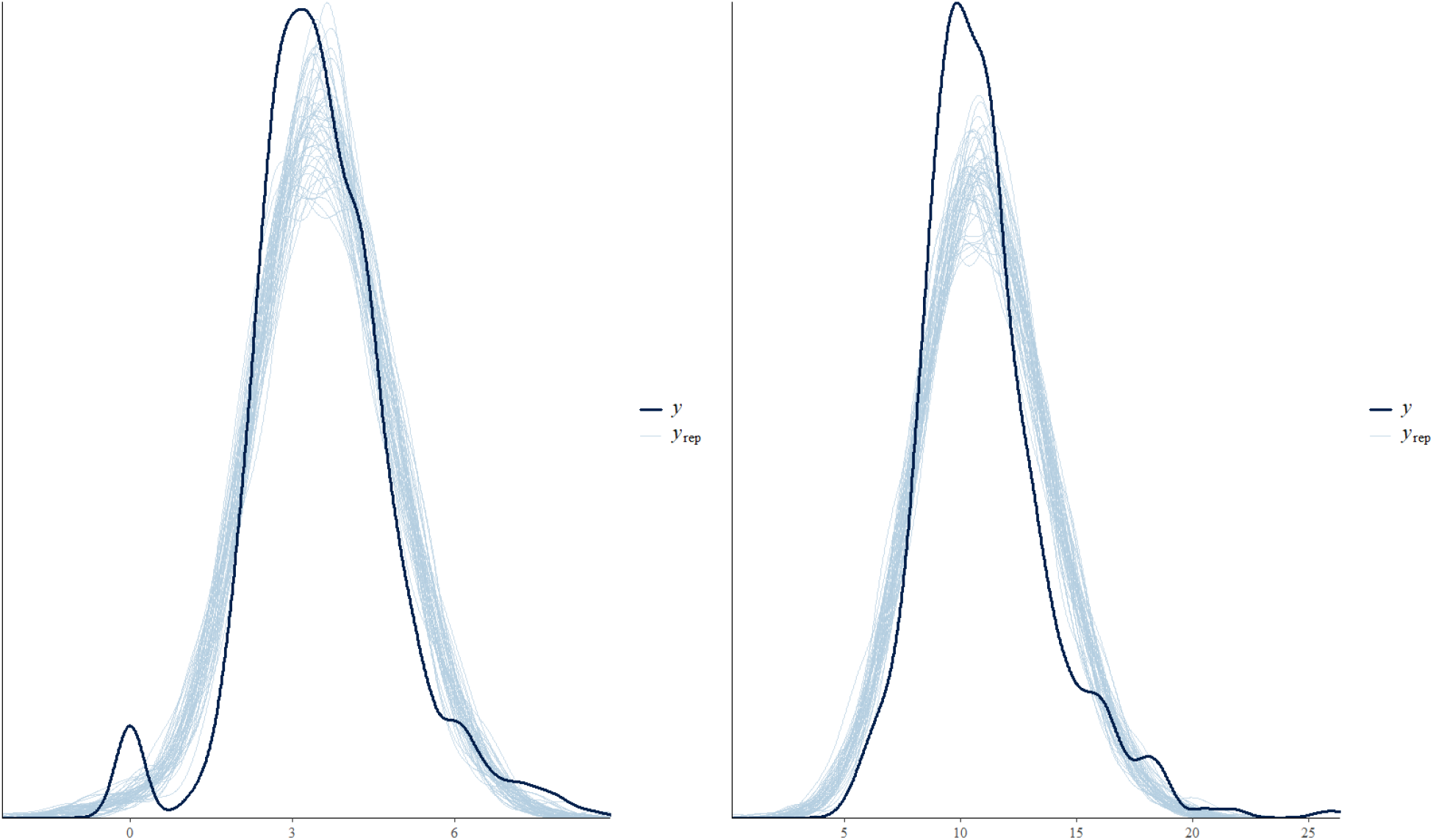
Comparison of posterior predictive distributions versus true distributions for LBW and PTB respectively. The true distribution is in dark blue. The posterior predictive distributions are in light blue. Substantial overlap and general matching of the shape of the distributions indicates serviceable model fit.

**Figure 6.**
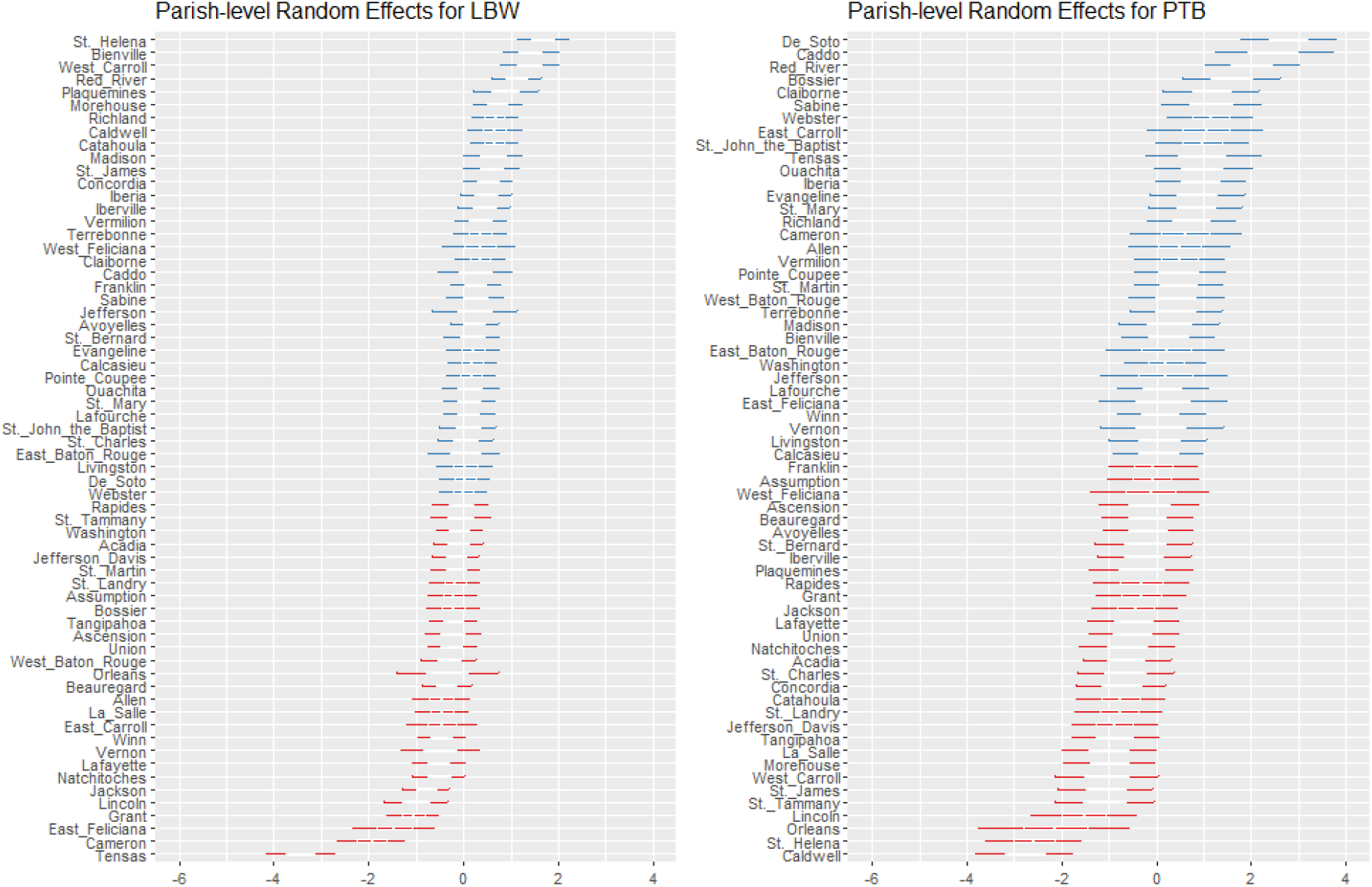
Parish-level random effects for LBW and PTB, respectively. They are sorted in decreasing order and colored by the directionality of their point estimates where a positive point estimate will be blue and a negative one will be read. They are presented alongside their 95% CI. The significant parish random effects for LBW are, in decreasing order, St. Helena, Bienville, West Carroll, Red River, Plaquemines, Morehouse, Richland, Caldwell, Catahoula, Jackson, Lincoln, Grant, East Feliciana, Cameron and Tensas. The significant parish random effects for PTB are, in decreasing order, De Soto, Caddo, Lincoln, Orleans, St. Helena, and Caldwell.

**Figure 7.**
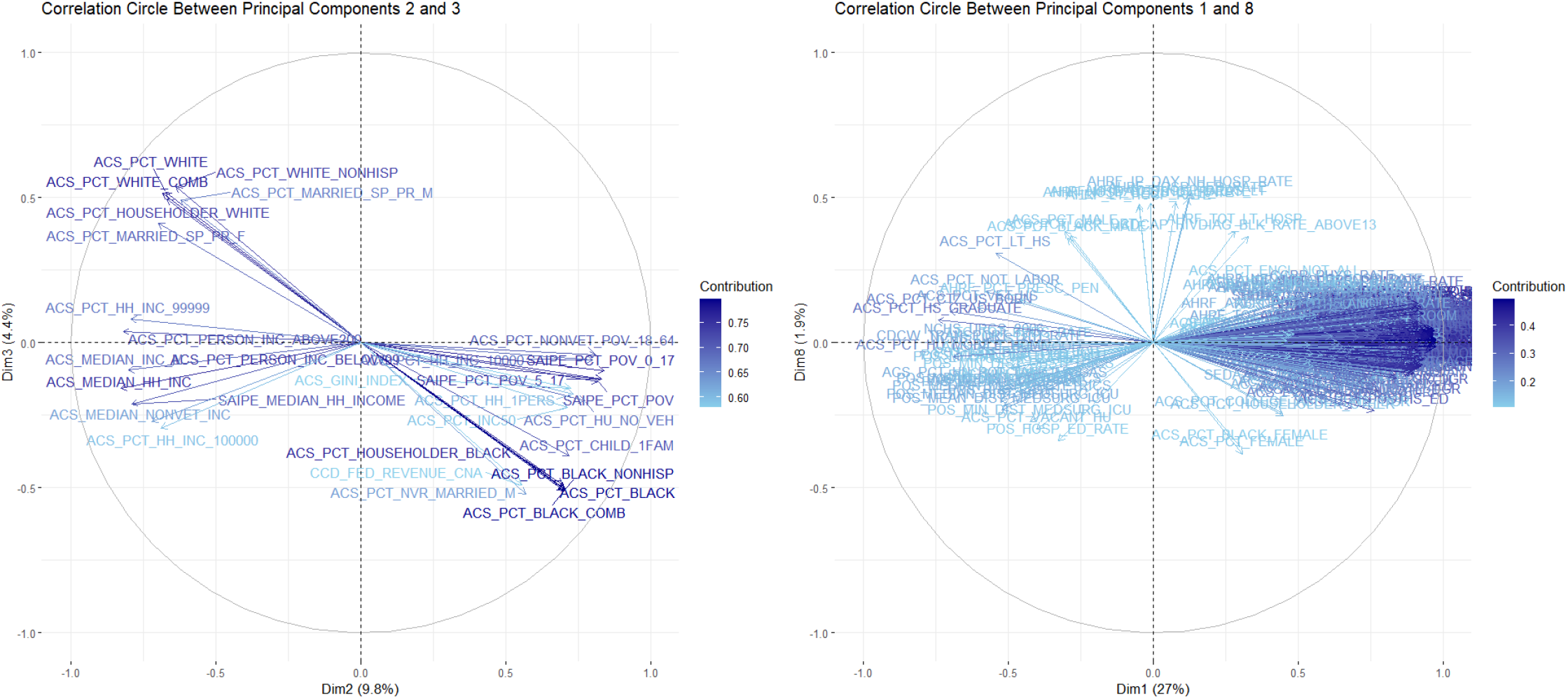
Correlation circles between principal components that were significant in at least one of our BLME models. The X, Y coordinates of each variable represent its correlations with each PC. The shade represents the importance of a variable’s contribution to each PC. The groups of variables represent correlation patterns between themselves. For example, the variable ACS_PCT_HOUSEHOLDER_BLACK (APHB) has coordinates .7, −5. Therefore, as the percentage of householders which self-report as Black and/or African American increases, PC 3 increases and PC 2 decreases. It is almost perfectly correlated with ACS_PCT_BLACK since they have nearly the same directionality and magnitude. It is nearly perfectly anticorrelated with ACS_PCT_WHITE since they have opposing directionality. It is also nearly uncorrelated with ACS_PCT_HH_INC_100000 since their directions are almost perpendicular.

**Figure 8.**
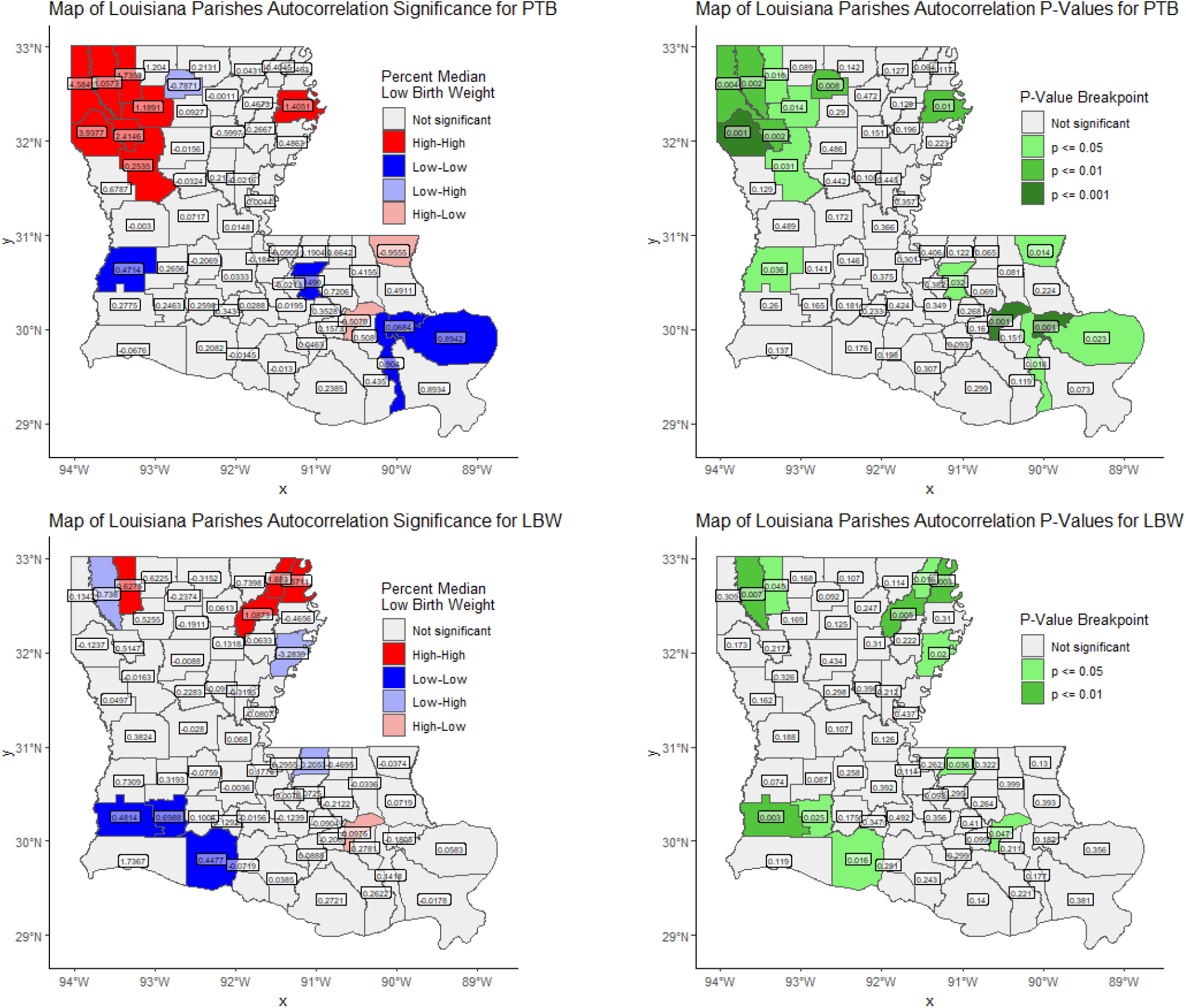
Chloropleth maps of the state of Louisiana colored by Local Moran’s I group and autocorrelation significance level, respectively. Parishes are overlayed with their Local Moran’s I statistic and p-value. Several clusters are identified here. notably St. John the Baptist Parish is classified as high-low on both maps. Additionally, Webster Parish is high-high on both maps. Finally, more low-low clusters show up in Southern Louisiana, while more high-high parishes appear in Northern Louisiana.

### Local Moran’s I

We utilized the queen neighbors setting to calculate the local indicators of spatial autocorrelation required by local Moran’s I for our polygons. The queen neighbors setting, when compared to the rook neighbors setting, has a higher distance tolerance between polygon centroids when determining whether they are neighbors. This allowed our analysis to be more lenient in terms of which parishes were considered neighbors, reflecting a wider network of interconnection between parishes. This analysis revealed sever clusters of spatial autocorrelation which can be seen in Figure 8. Several parishes were classified as high-high meaning that they themselves have a high value of the respective ABO while being surrounded by other parishes with high values. All of these were in Northern Louisiana which is a more rural area of Louisiana. At the same time, low-low clusters were identified in Southern Louisiana. Southern Louisiana contains most of the major cities within the states. This indicates an important component of rurality vs urbanicity in the incidence of ABO. Some parishes were identified as outliers in terms of their surroundings, granting them high-low or low-high status. This means that they have higher or lower values of ABO incidence than average, respectively, in comparison to their surroundings which accordingly are lower or higher than the mean. Figure 8 also illustrates the respective p-value for each parish which is useful to ascertain the certainty of their classification.

### Combined Analysis

We selected the best predictors for both LBW and PTB from the BLME models based on their 95% confidence regions. These were PC2 and PC3 for LBW, and PC1, PC2 and PC8 for PTB, respectively. Then we projected all 12 observations per parish onto 2-dimensional or 3-dimensional principal component space, as needed. Following this we overlayed them with their calculated local Moran’s I label. Examining Figure 9 and Figure 10 allowed us to find patterns of similarities between parishes assigned similar labels. Among these, in Figure 9 we can appreciate a hyperplane approximately where y=0. No high-high clusters exist to the left of this line, the same can be said for low-low parishes on the right side. Thus, this provides additional evidence for the importance of the variables accounted for in PC 2 as determining factors in LBW incidence rate at the parish level. In Figure 10, we can see that the 4 most populous parishes in Louisiana break out from the others. Mostly this solidifies the relationship between urbanicity and lowered rates of preterm birth since all but one for these high population parishes were classified as low-low. Of these, Caddo Parish is the only one to be classified as high-high. Thereby being an exception to this trend. We hypothesize this to be because of two major, intersecting factors: larger distance between Shreveport and other major cities in Louisiana, and more frequent exposure to risk factors not considered in our models such as occurrence of teenage and young adult pregnancies carried to delivery and increased rates of drug use. In fact, from 2014 to 2023 Caddo Parish has consistently shown rates of births by mothers below the age of 20 that are higher than those of Orleans Parish, Jefferson Parish, and East Baton Rouge Parish despite its lower overall population and births per year.[17]

**Figure 9.**
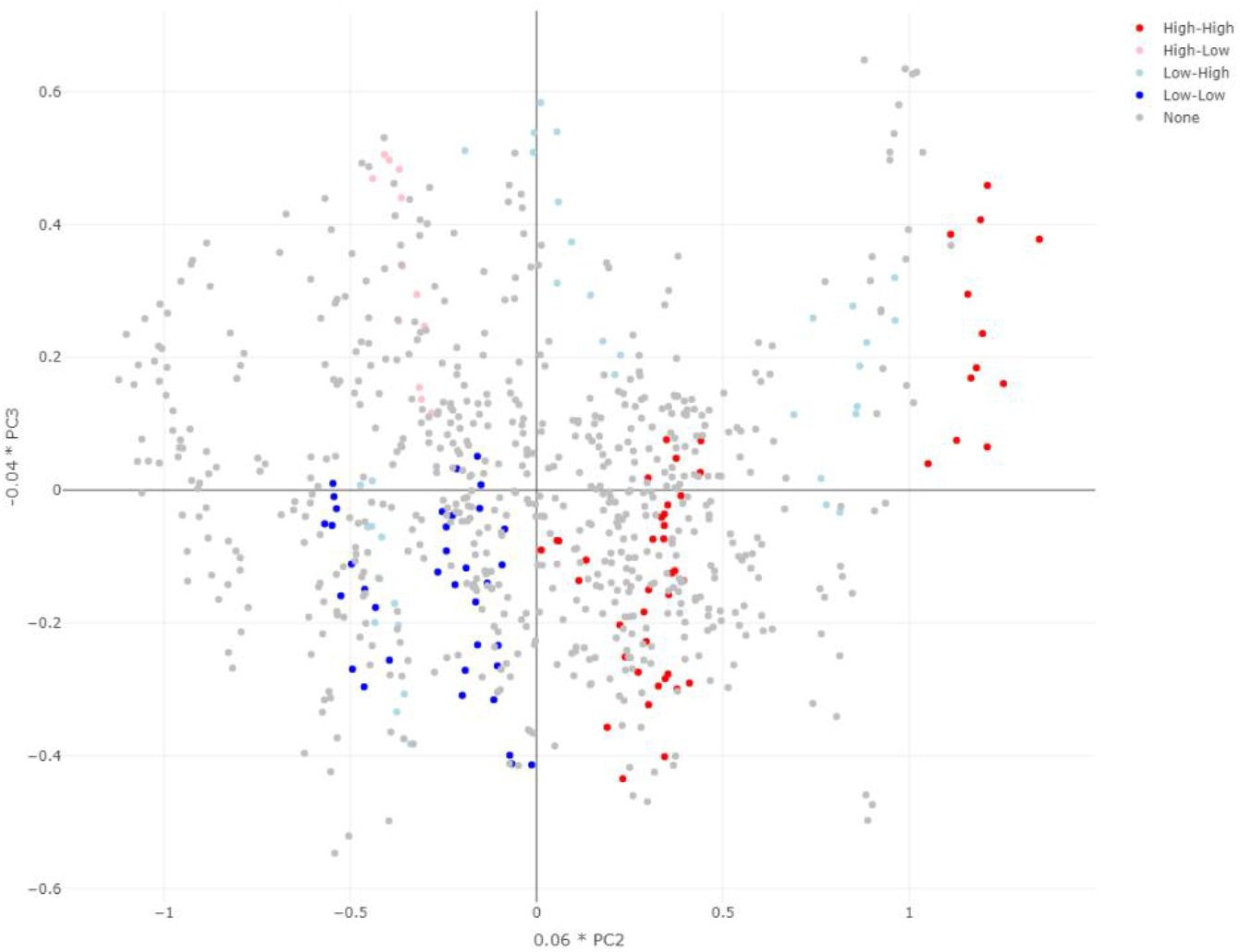
2-Dimensional principal component space plot overlayed with local Moran’s I labels for LBW.

**Figure 10.**
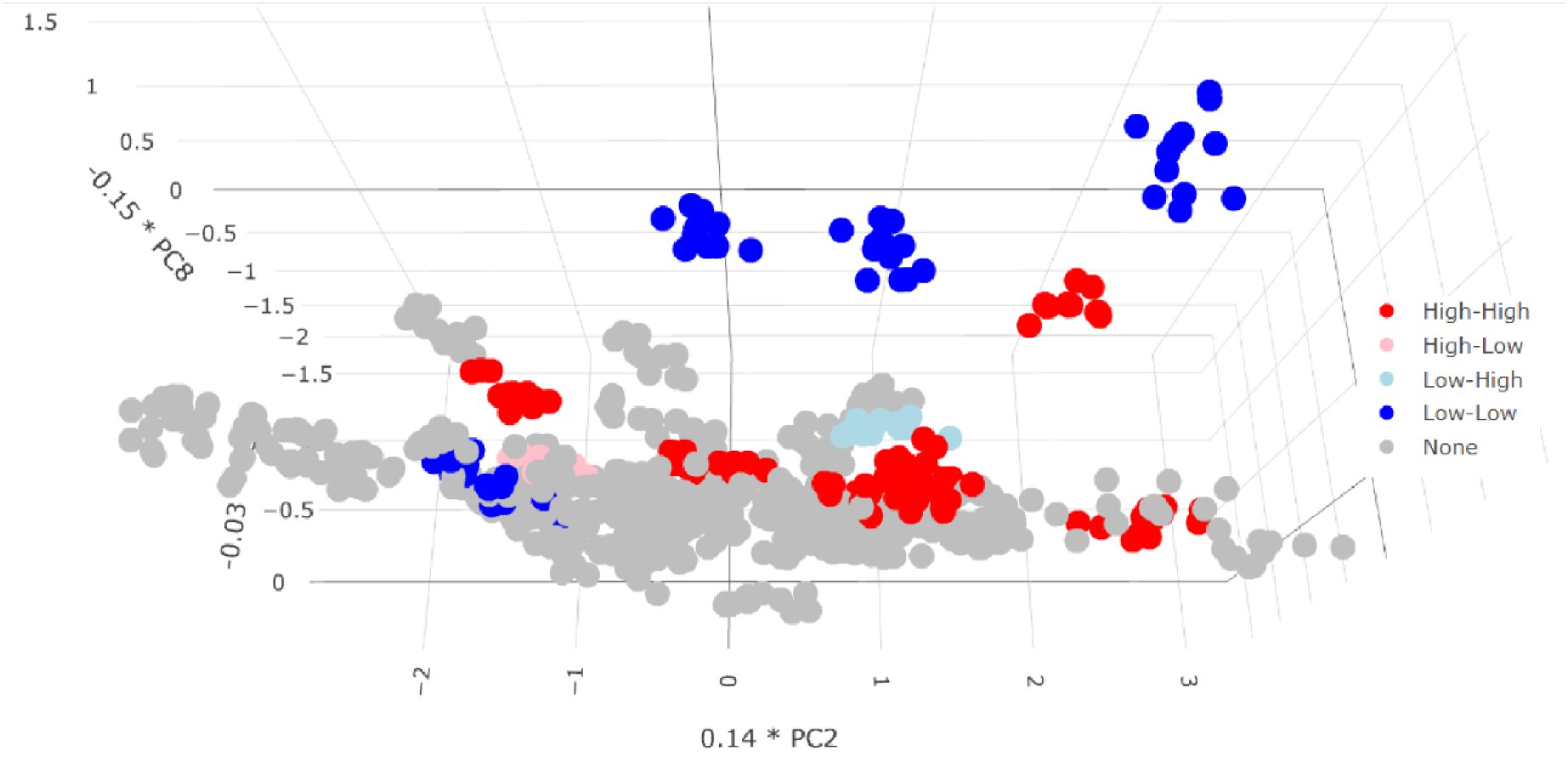
3-Dimensional principal component plot overlayed with local Moran’s I labels for PTB. Parishes with higher population density like Orleans, Caddo, Jefferson, and East Baton Rouge occupy the upper part of the space. Of these all, except Caddo, are classified as low-low while Caddo is classified as high-high. The two low parishes aside from the major cities are Lincoln, Beauregard, and St. Bernard. Of these, St. Bernard is proximal to both Orleans and Jefferson

## Discussion

### Key Findings

Our models indicate that there are significant spatial and SDoH-linked effects on the incidence of ABO in Louisiana. The spatial effects include parishes that significantly modify the prediction of ABO rates as well as clusters of disproportionately high and low rates of ABO. The correlation circles reveal that the social determinants of health that contribute most to the differential incidence of PTB in Louisiana are previously reported factors, such as populational racial composition, economic prosperity, and household size. [2,6] For LBW, the interpretation is more challenging. The fact that PC1 is significantly associated with LBW incidence despite our LBW measurements being normalized in terms of populational size is particularly vexing. We hypothesize that this reaffirms previously documented associations between higher populational density lowering the incidence of LBW.[18] PC2 being significant, as with PTB, underscores the idea that racial composition and economic prosperity within a parish is informative of the rates of incidence for LBW.[6] Finally, because PC8 significantly predicts LBW we find that increased healthcare utilization decreases the rate of LBW incidence in a parish. On the other hand, we found no statistically significant associations between years in our dataset. This indicates that the rates of ABO incidence in Louisiana have not significantly changed over time. While they did not worsen in the timeframe we examined, there was also no significant improvement. This is especially interesting to note when there were policy-based initiatives within the time frame of our study aimed at improving maternal and child health. Particularly, the Louisiana Birth Outcomes Initiative was active between 2010 and 2014 and sought to reduce preterm births. [19, 20] Our results indicate that this initiative did not move the needle on a state level, possibly because of their lack of focus on health disparities stemming from underlying SDoH. [20]

Another important finding of our analysis is the identification of clusters of autocorrelated parishes in terms of ABO. Particularly high-high parishes represent clusters of parishes which are especially vulnerable to ABO. These high-high parishes were found exclusively in North Louisiana. We also identified high-low parishes which, in isolation, have significantly higher rates of ABO than their neighbors. The only parish which appears as high-low in both plots is St. John the Baptist Parish. St. John the Baptist Parish is, notably, one of the parishes within Louisiana that forms Louisiana’s Industrial Corridor. St. John specifically has been greatly affected by the environmental conditions caused by surrounding chemical plants, being classified as the county with the highest rate of cancer in the U.S.[21, 22] Here, it is evidently also disproportionately affected by ABO when accounting for its neighboring parishes. These neighboring parishes compose a Low-Low cluster in terms of PTB. The other cluster of Low-Low ABO we identified was for LBW. In contrast to the High-High parishes, we found Low-Low parishes exclusively in Southern Louisiana.

### Limitations

The main limitation of the BLME-based modelling part of this work is that the data available represents an interval of time that may not be representative of the full history of ABO in Louisiana. With a larger range of time, we could calculate better imputed values to handle missingness, incorporate additional SDoH, and potentially identify time-based effects. For example, this work does not capture the effects of the SARS-CoV-2 pandemic on the incidence of ABO due to the range of years which were available at the time of data analysis. Meanwhile, the main limitation of the results of the identification of spatial clusters is that Louisiana does not exist in a geographical vacuum. That is, there are adjacent geographies in the states of Texas, Arkansas, and Mississippi whose values of ABO incidence were not considered. Considering the ABO incidence rates for these geographies could alter cluster assignment, particularly for parishes on the northern, western, and eastern borders of Louisiana.

## Conclusion

Our BLME models had perfect R-hat statistics, reasonable similarity between the posterior predictive distributions and the true distributions of our ABO rates, and serviceable Bayesian R^2^ metrics. Taking this into consideration we found no reason to conclude that our models do not fit the data well. From our BLME models, we found evidence of associations between parish and SDoH-derived principal components. From those principal components, the major factors for PTB seem to be differences in racial composition and economic prosperity. While for LBW racial composition and economic prosperity were also important factors, differences in population density and healthcare utilization. We also reported various parishes with spatially correlated measures of ABO. High ABO rate parishes surrounded by other high ABO rate parishes seem to be more common in North Louisiana. Low ABO rate parishes surrounded by other low ABO rate parishes were found to be more common in South Louisiana. Future research in this area could assess why high-high clusters of ABO in Louisiana are strictly in North Louisiana and Low-Low clusters only occur in South Louisiana. Some parishes with discordant measures of ABO rates were identified. Particularly, St. John the Baptist Parish, one of the parishes within Louisiana’s Industrial Corridor, was identified as significantly higher than its neighbors as well as identifying clusters of high ABO rates in Northern Louisiana. No year-related associations were identified, indicating no significant change in ABO rates over time in Louisiana even though there were policies aimed at addressing ABO on a state level. In short, we were able to predict the rates of ABO in Louisiana well. We identified no significant temporal associations with the incidence of ABOs in Louisiana, however there were significant place-based and SDoH-related associations. In addition, the objective selection of SDoH data from a large dataset largely replicates previously documented associations with the rates of ABO, especially parish-level racial composition, population density, healthcare utilization, and economic attainment.[2,6,18]

## Equations

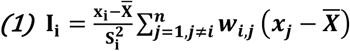

Equation 1. Calculation of Local Moran’s I statistic for the *i*^*th*^ data point in a data set with n observations. Here, *x*_*i*_ is the value of the variable being examined for the *i*^*th*^ observation, 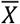is the mean value for that variable, 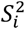is the variance of the variable in all observations except I which is further explained in equation 2, *w*_*i,j*_is a measure of spatial weight between the *i*^*th*^ and *j*^*th*^ observations.

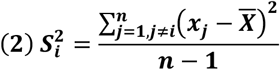

Equation 2. Calculation of spatial variance measure used in the calculation of Local Moran’s I for the *i*^*th*^ data point.

## Data Availability

The datasets used in this work are available from the Agency of Health Research and Quality and the Louisiana Department of Health.

https://healthdata.ldh.la.gov/

https://www.ahrq.gov/data/innovations/clh-data.html

## Funding acknowledgement

Research reported in this publication was partly supported by the National Institute Of General Medical Sciences of the National Institutes of Health under Award Number P20GM152305. The content is solely the responsibility of the authors and does not necessarily represent the official views of the National Institutes of Health.

